# Estimation of the final size of the second phase of the coronavirus COVID 19 epidemic by the logistic model

**DOI:** 10.1101/2020.03.11.20024901

**Authors:** Milan Batista

## Abstract

In the note, the logistic growth regression model is used for the estimation of the final size and its peak time of the coronavirus epidemic in China, South Korea, and the rest of the World.

## 1 Introduction

In the previous article [1], we try to estimate the final size of the epidemic for the whole World using the logistic model and SIR model. The estimation was about 83000 cases. Both models show that the outbreak is moderating; however, new data showed a linear upward trend. It turns out that the epidemy in China was slowing but is begin to spread elsewhere in the World.

In this note, we will give forecasting epidemic size for China, South Korea, and the rest of the World and daily predictions using the logistic model. Full daily reports for China, Germany, Iran, Italy, Slovenia, South Korea, Spain and counties outside of China generated are available at https://www.researchgate.net/publication/339912313_Forecasting_of_final_COVID-19_epidemic_size

The MATLAB program *fitVirus* used for calculations is freely available from https://www.mathworks.com/matlabcentral/fileexchange/74411-fitvirus

We note that logistic models give similar results as the SIR model (at least for the case of China and South Korea). However, the logistic model is given by explicit formula and is thus much simpler for regression analysis than the SIR model, where one must on each optimization step solve a system of ordinary differential equations. (One may, however, use approximate solution and thus obtain four-parameter problem which can be very sensitive to initial guess). Yet, the logistics model has its drawbacks as the epidemic approaches its final stage: the actual number of cases may be slightly larger than that predicted by the logistics model. If the actual number of cases begins to exceed the predicted end-state systematically, then a second phase of the epidemic is likely to occur, and the model will no longer be applicable.

## 2 Logistic grow model

In mathematical epidemiology, when one uses a phenomenological approach, the epidemic dynamics can be described by the following variant of logistic growth model [2-5]

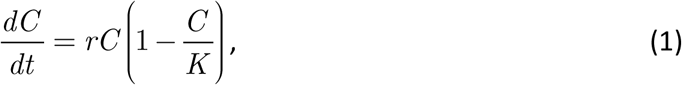

where *C* is an accumulated number of cases, *r* > 0 infection rate, and *K* > 0 is the final epidemic size. If *C* (0) =*C* _0_ > 0 is the initial number of cases then the solution of (1) is

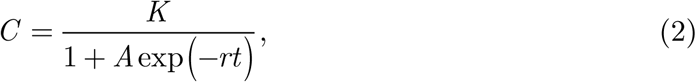

Where 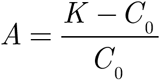 When *t* ⟪ 1, assuming *K* ⟫ *C* _0_, and therefore *A* ⟫ 1 we have the natural growth

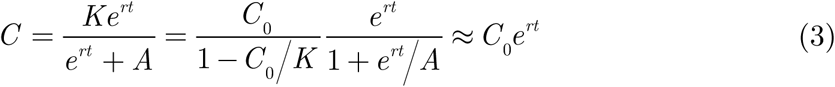

When *t* → ∞ the number of cases follows the Weibull function

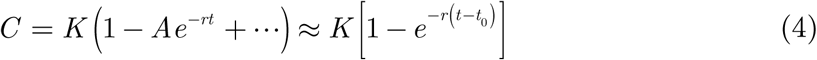

The growth rate 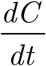 reaches its maximum when 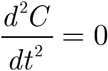. From this condition, we obtain that the growth rate peak occurs in time time

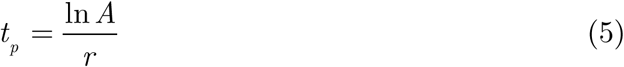

At this time the number of cases is

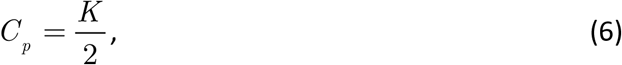

and the growth rate is

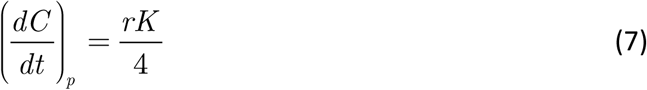

To answer the question about doubling time Δ*t*, i.e., the time takes to double the number of cases we solve *C* (*t* + Δ*t*) =2*C* (*t*) for Δ*t*. Result is

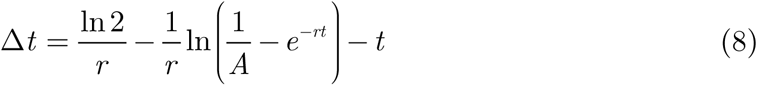

The first term represents initial exponential growth, then Δ*t* increases with *t*. When 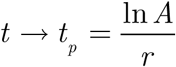, i.e., when *C* → *K/*2, then Δ*t* → ∞. For *C* ≥ *K/*2 doubling time lost its meaning.

Now, if *C*_1_,*C*_2_,…,*C*_*n*_ are the number of cases at times *t*_1_,*t*_2_,…,*t*_*n*_, then the final size predictions of the epidemic based on these data are *K*_1_, *K*_2_,…, *K*_*n*_. When convergence is achieved, then one may try to predict the final epidemic size by iterated Shanks transformation [6]

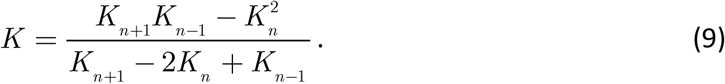

There is no natural law or process behind this transformation; therefore, it must be used with some care. In particular, the calculated limit is useless if *K* < *C*_*n*_, i.e., it is below the current data.

The logistic model (2) contains three parameters: *K, r*, and *A*, which should be determined by regression analysis. Because the model is nonlinear, some care should be taken for initial guess. First of all, in the early stage, the logistic curve follows an exponential growth curve (3), so the estimation of *K* is practically impossible. With enough data, the initial guess can be obtain in the following way. Expressing *t* from (2) and use three equidistant data point yield the following system of three equations:

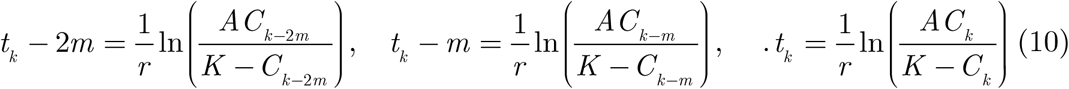

This system has a solution [7]

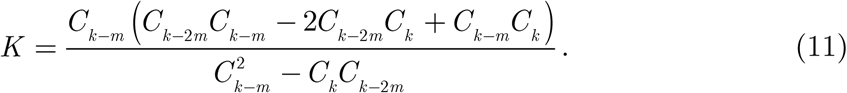

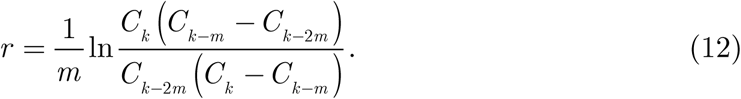

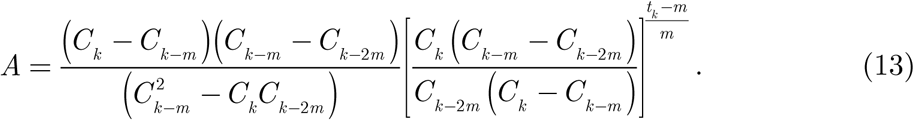

The solution is acceptable when all the unknowns are positive.

Formulas (10),(11),(12) are used to calculate the initial approximation in the *fitVirus* program. For practical calculation, we take the first, the middle, and the last data point. If this calculation fails, we consider regression analysis as questionable. Final values of the parameters *K, r*, and *A* are then calculated by least-square fit using the MATLAB functions *lsqcurvefit* and *fitnlm*.

Before we proceed, we for convenience, introduce the following epidemy phases (see Fig 2):

**Figure 1.**
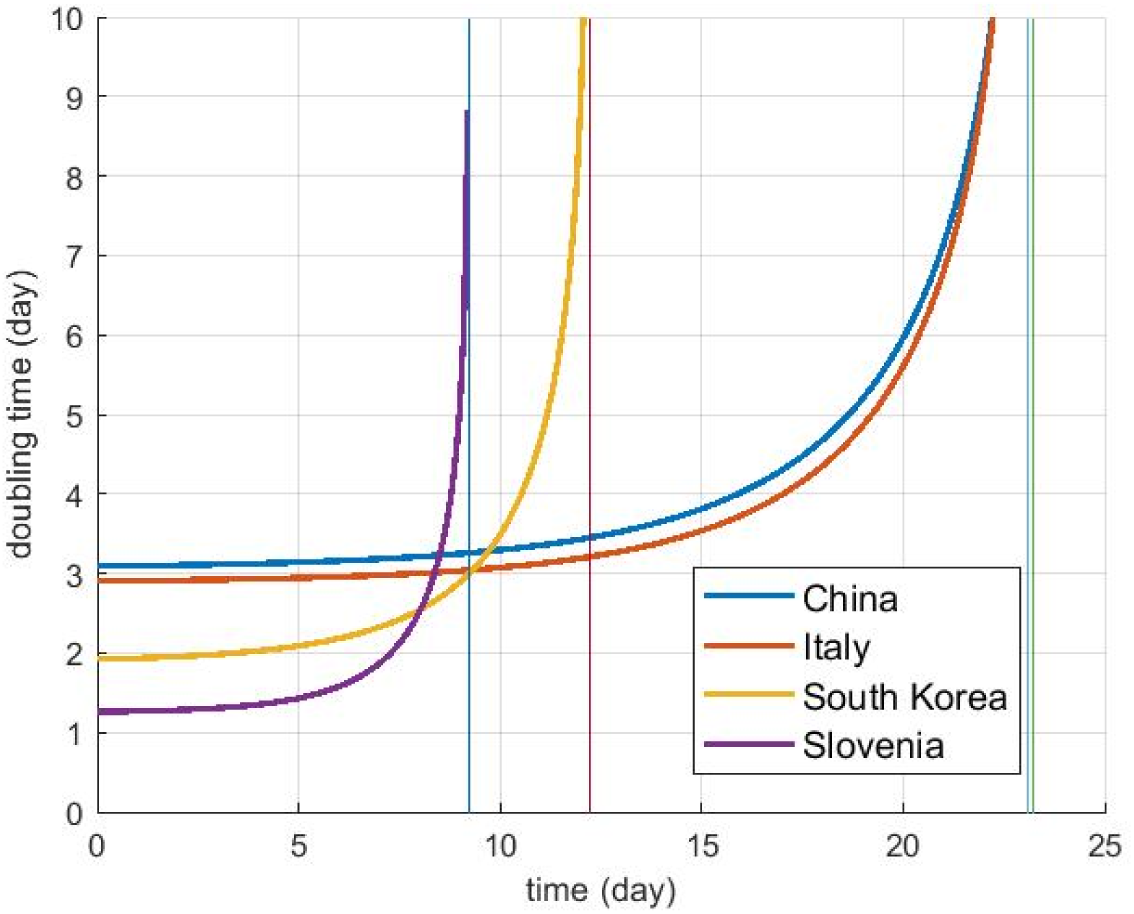
Doubling time (data up to 14 mar 2020)

**Figure 2.**
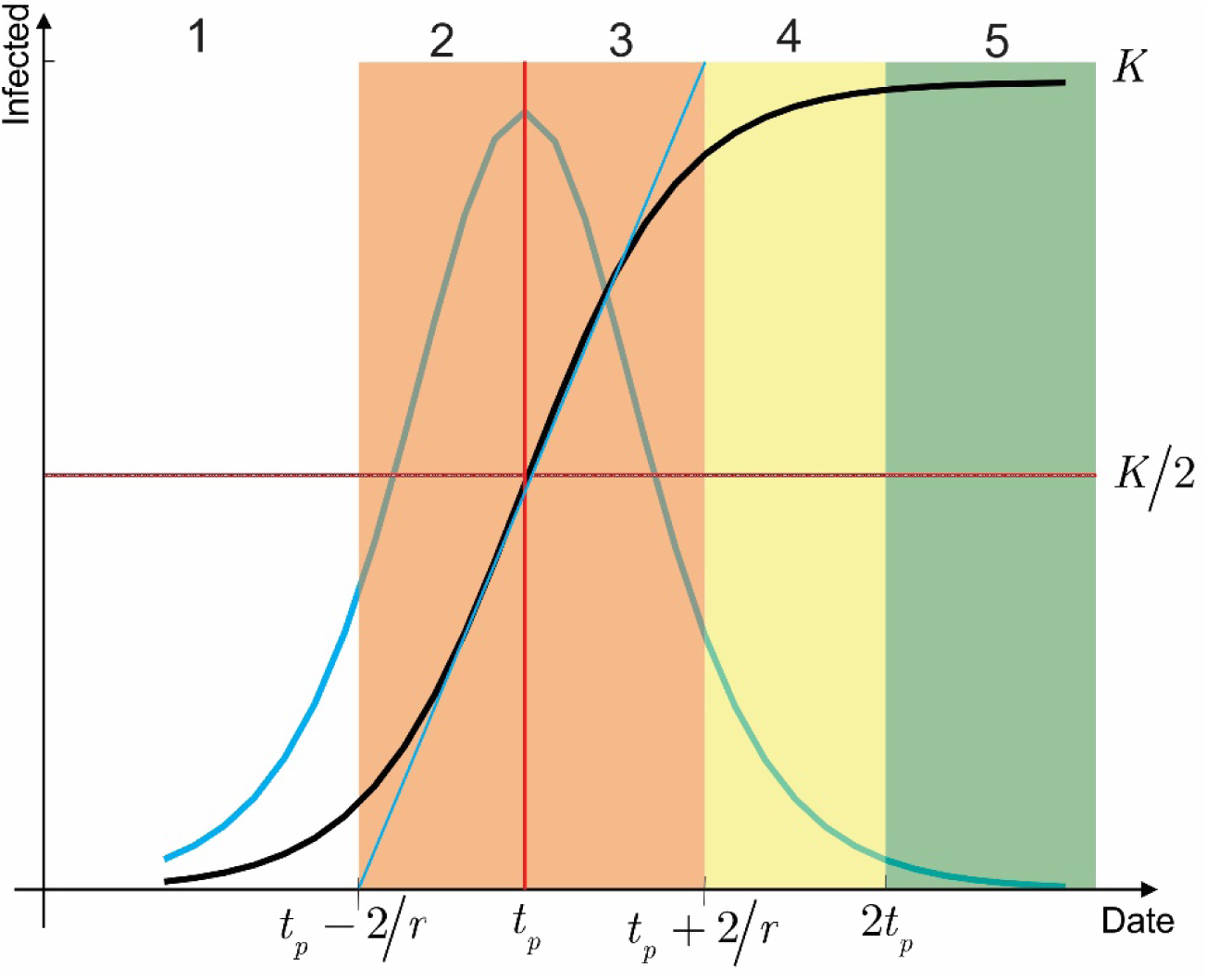
Epidemic phases.

1. Phase 1 – exponential growth (lag phase, slow growth) : *t*_*p*_ < *t* − 2/*r*

2. Phase 2 – fast growth (positive growth phase, acceleration phase) to an epidemy turning point: *t*_*p*_ − 2/ *r* < *t* < *t*_*p*_

3. Phase 3 – fast growth to steady-state (negative growth phase, deceleration phase): *t*_*p*_ < *t* < *t*_*p*_ + 2/*r*

4. Phase 4-steady-state (transition phase, slow growth, asymptotic): *t*_*p*_ + 2 /*r* < *t* < 2*t*_*p*_

5. Phase 5 – steady) ending phase (plateau stage): *t* > 2*t*_*p*_

The duration of the fast-growing period is thus equal to *τ* =4/ *r*. We note that the names of the phases are not standard, and are arbitrarily chosen.

## 3 Results

### 3.1 *China* (11.Mar 2020)

On the base of available data, one can predict that the final size of coronavirus epidemy in China using the logistic model will be approximately 81 000 ± 500 cases (Table 1) and that the peak of the epidemic was on 8 Feb 2020 (Table 2). It seems that the epidemic in China is in the ending stage (Fig 3, Fig 4).

**Table 1.** Estimated logistic model parameters for China (data up to 11.Mar 2020)

|  | Estimate | SE | tStat | pValue |
| --- | --- | --- | --- | --- |
| **K** | **80772** | 489.04 | 165.16 | 2.0205e-72 |
| r | 0.22568 | 0.0062941 | 35.855 | 2.4094e-38 |
| A | 191.23 | 26.978 | 7.0882 | 3.5795e-09 |

Number of observations: 55, Error degrees of freedom: 52 Root Mean Squared Error: 1.9e+03 R-Squared: 0.997, Adjusted R-Squared 0.997 F-statistic vs. zero model: 1.61e+04, p-value = 4.02e-77

**Table 2.**
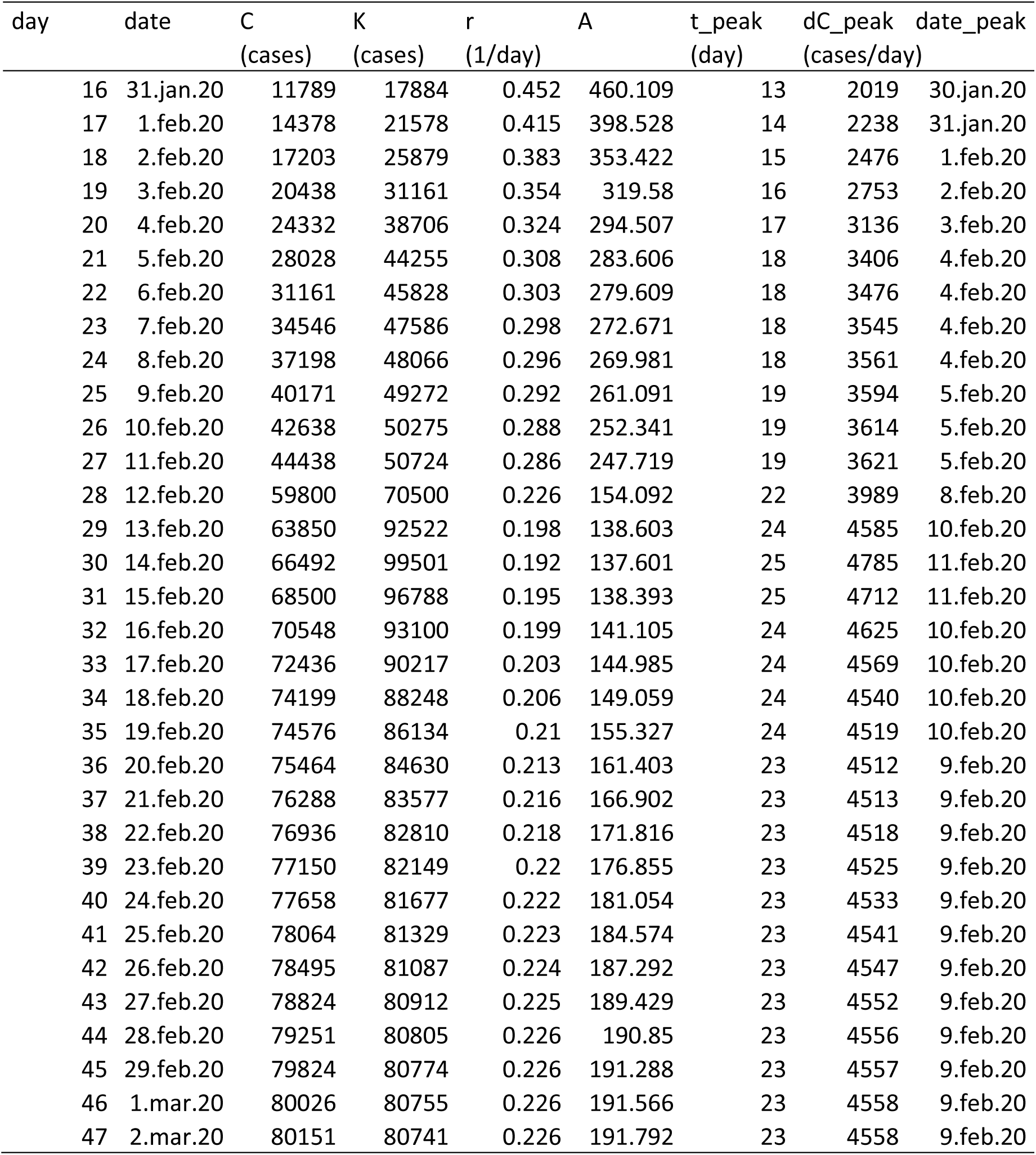
Results of daily logistic regression for China (data from 4.Mar 2020)

**Figure 3.**
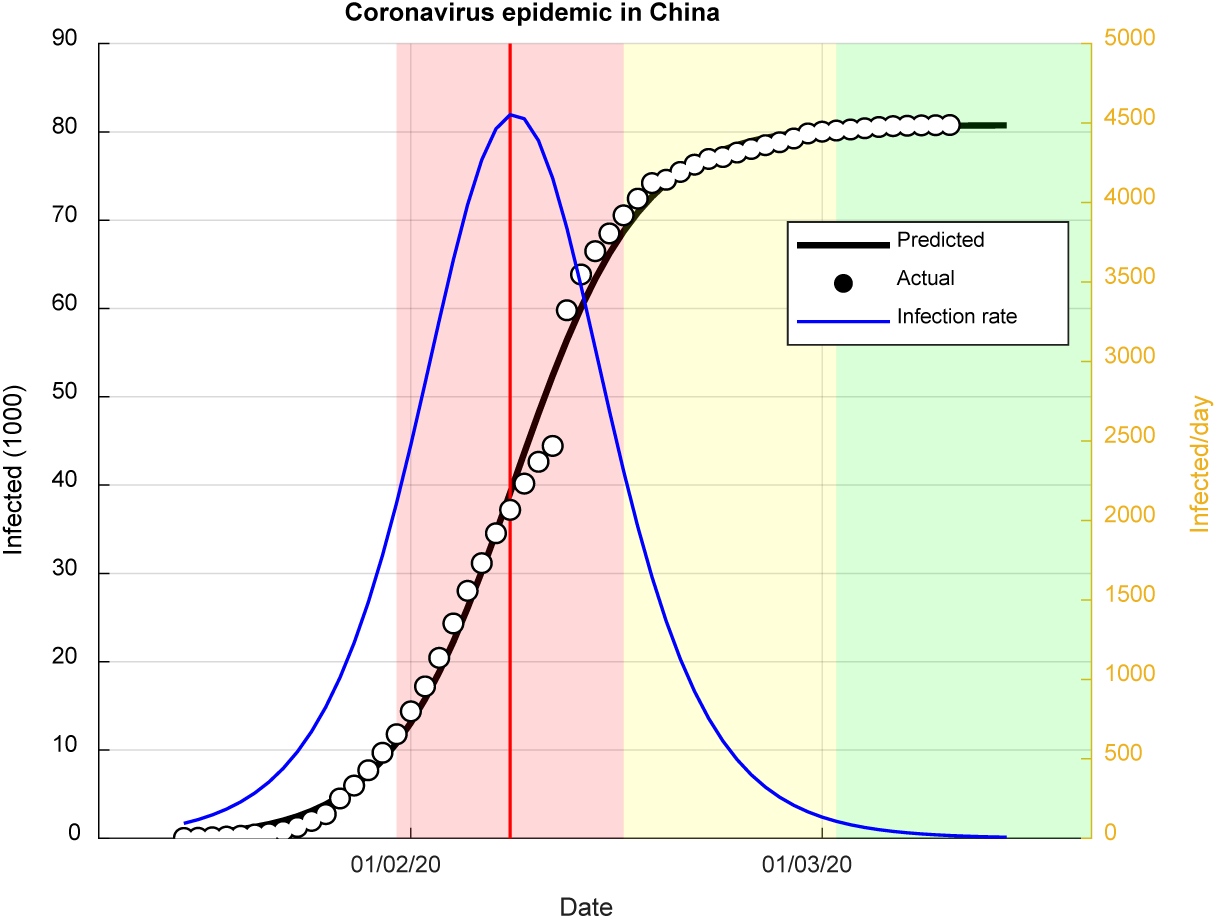
Predicted evaluation of coronavirus epidemic in China.

**Figure 4.**
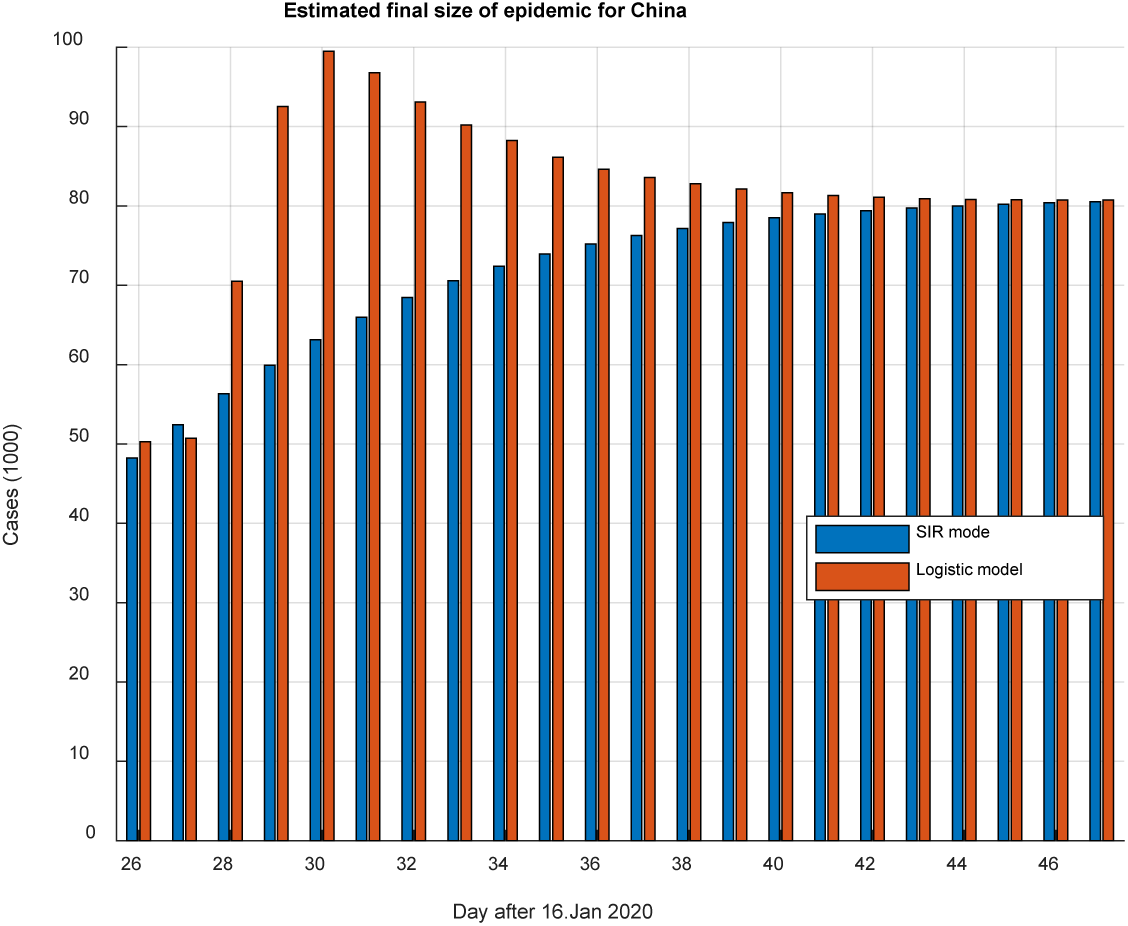
Predicted final size of coronavirus epidemic in China (prediction from 2.Mar 2020)

The short-term forecasting is given in Table 3 where we see that the discrepancy of actual and forecasted number of cases is within 2%. However, actual and predicted daily new cases are scattered and vary between 13% to 300%. On 7 Mar 2020, the actual number of cases was 80695, and the daily number of cases was 44. Prediction in Table 3 is cumulative 80588 cases and 39 daily cases. The errors are 0.1% and 11%, respectively.

**Table 3.** Short-term forecasting for China

| Day | Date | Actual | Predicted | Error % | Daily actual | Daily predicted | Error % |
| --- | --- | --- | --- | --- | --- | --- | --- |
| 44 | 28.feb.20 | 79251 | 79814 | 0.945 | 427 | 232 | 45.667 |
| 45 | 29.feb.20 | 79824 | 80000 | 0.407 | 573 | 186 | 67.539 |
| 46 | 1.mar.20 | 80026 | 80149 | 0.302 | 202 | 149 | 26.238 |
| 47 | 2.mar.20 | 80151 | 80268 | 0.265 | 125 | 119 | 4.8 |
| 48 | 3.mar.20 | - | 80363 | - | - | 95 |  |
| 49 | 4.mar.20 | - | 80439 | - | - | 76 |  |
| 50 | 5.mar.20 | - | 80500 | - | - | 61 |  |
| 51 | 6.mar.20 | - | 80549 | - | - | 49 |  |
| 52 | 7.mar.20 | - | 80588 | - | - | 39 |  |

### 3.2 *South Korea* (11.Mar 2020)

On the base of available data, one can predict that the final size of coronavirus epidemy in of South Korea using the logistic model will be approximately 8050 ±70 cases (Fig 5, Table 4) and that the peak of the epidemic was on 1 Mar 2020. The epidemic in South Korea appears to be in the steady-state transition phase. These figures were already predicted on 4. Mar 2020 (Table 5), i.e., the prediction was approximately 7500 to 8500 cases and that the peak will be around 2 Mar.

**Table 4.** Estimated logistic model parameters for South Korea up to 13.Mar 2020

|  | Estimate | SE | tStat | pValue |
| --- | --- | --- | --- | --- |
| K | 8046.5 | 71.784 | 112.09 | 7.8279e-32 |
| r | 0.36657 | 0.010259 | 35.733 | 5.581e-21 |
| A | 87.97 | 9.8747 | 8.9087 | 9.4612e-09 |

Number of observations: 25, Error degrees of freedom: 22 Root Mean Squared Error: 129 R-Squared: 0.998, Adjusted R-Squared 0.998 F-statistic vs. zero model: 1.26e+04, p-value = 1.03e-35

**Table 5.** Results of daily logistic regression for South Korea

| day | date | C<br>(cases) | K<br>(cases) | r<br>(1/day) | A | t_peak<br>(day) | dC_peak<br>(cases/day) | date_peak |
| --- | --- | --- | --- | --- | --- | --- | --- | --- |
| 7 | 24.feb.20 | 833 | 1105 | 0.824 | 47.332 | 4 | 227 | 23.feb.20 |
| 8 | 25.feb.20 | 977 | 1133 | 0.808 | 46.526 | 4 | 228 | 23.feb.20 |
| 9 | 26.feb.20 | 1261 | 1508 | 0.633 | 37.083 | 5 | 238 | 24.feb.20 |
| 10 | 27.feb.20 | 1766 | 3355 | 0.426 | 44.537 | 8 | 357 | 27.feb.20 |
| 11 | 28.feb.20 | 2337 | 7021 | 0.363 | 76.656 | 11 | 636 | 1.mar.20 |
| 12 | 29.feb.20 | 3150 | 17145 | 0.328 | 165.531 | 15 | 1405 | 5.mar.20 |
| 13 | 1.mar.20 | 3736 | 8639 | 0.359 | 95.534 | 12 | 775 | 2.mar.20 |
| 14 | 2.mar.20 | 4335 | 7214 | 0.378 | 88.434 | 11 | 682 | 1.mar.20 |

**Table 6.**
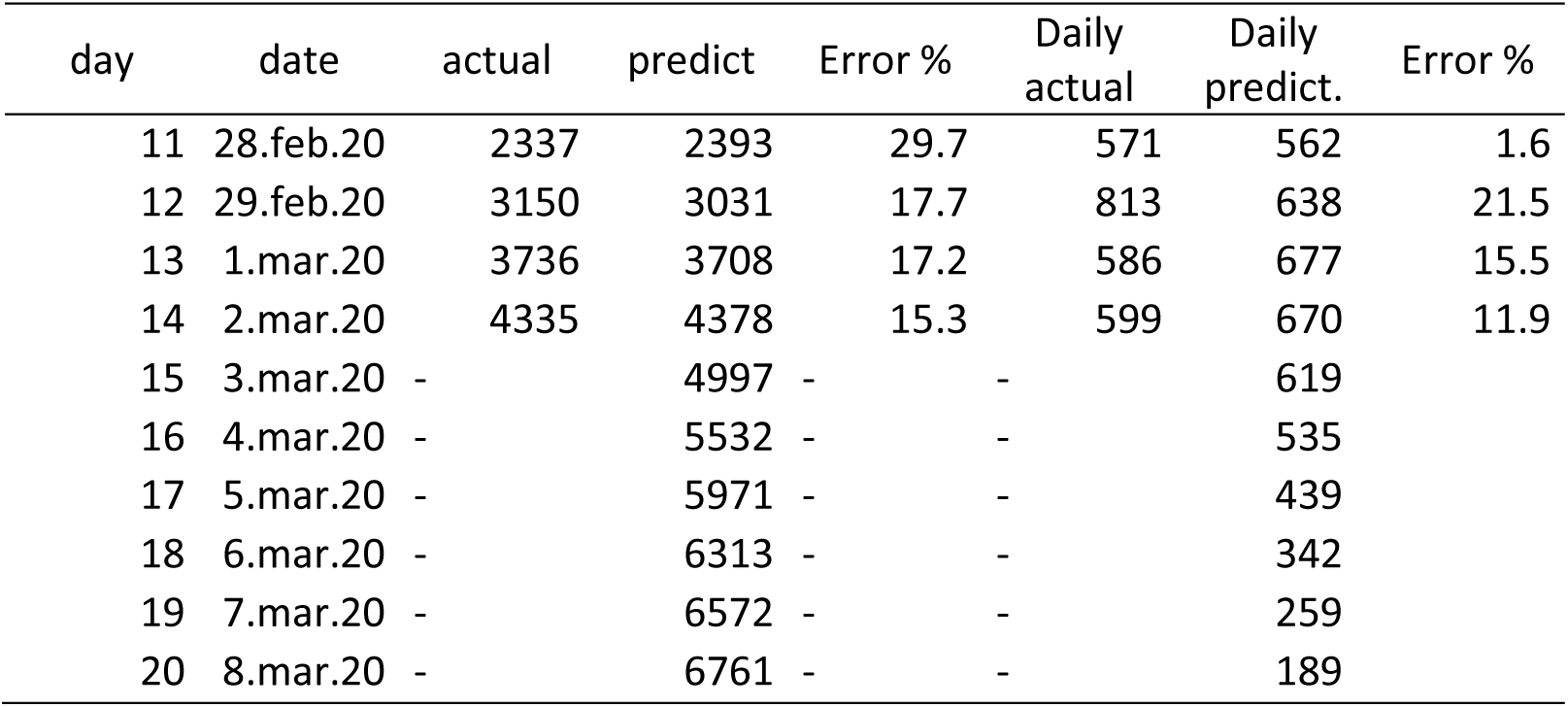
Short-term forecasting for South Korea

| day | date | actual | predict | Error % | Daily<br>actual | Daily<br>predict. | Error % |
| --- | --- | --- | --- | --- | --- | --- | --- |
| 11 | 28.feb.20 | 2337 | 2393 | 29.7 | 571 | 562 | 1.6 |
| 12 | 29.feb.20 | 3150 | 3031 | 17.7 | 813 | 638 | 21.5 |
| 13 | 1.mar.20 | 3736 | 3708 | 17.2 | 586 | 677 | 15.5 |
| 14 | 2.mar.20 | 4335 | 4378 | 15.3 | 599 | 670 | 11.9 |
| 15 | 3.mar.20 | - | 4997 | - | - | 619 | - |
| 16 | 4.mar.20 | - | 5532 | - | - | 535 | - |
| 17 | 5.mar.20 | - | 5971 | - | - | 439 | - |
| 18 | 6.mar.20 | - | 6313 | - | - | 342 | - |
| 19 | 7.mar.20 | - | 6572 | - | - | 259 | - |
| 20 | 8.mar.20 | - | 6761 | - | - | 189 | - |

**Figure 5.**
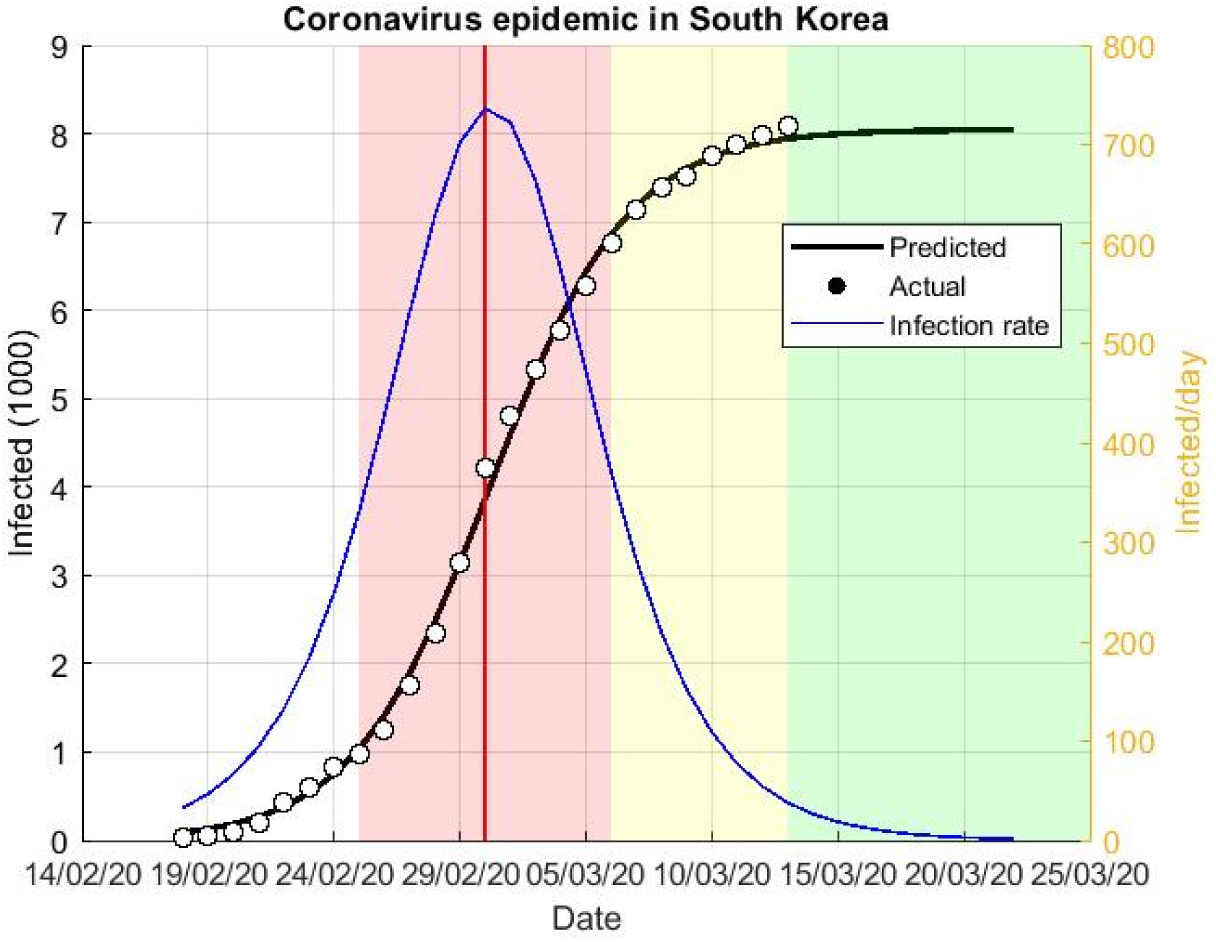
Predicted evaluation of coronavirus epidemic in South Korea (13.Mar 2020)

On 7 Mar 2020, the actual number of cases was 7134, and the daily number of cases was 367. Prediction in Table 5 is cumulative 6572 cases and 259 daily cases. The errors are 8 % and 30%, respectively.

#### 3.3. Rest of the World

The comparison of the predicted final sizes is shown in the graph in Figure 6.Based on the data from 11. Mar 2020, a very rough estimate indicates that the number of cases will be about 90000 (Fig 7); data from 13. Mar 2020 rise this number to 380 000. However, it is an early-stage epidemic, so these estimates are very questionable and will be changed with new data.

**Figure 6.**
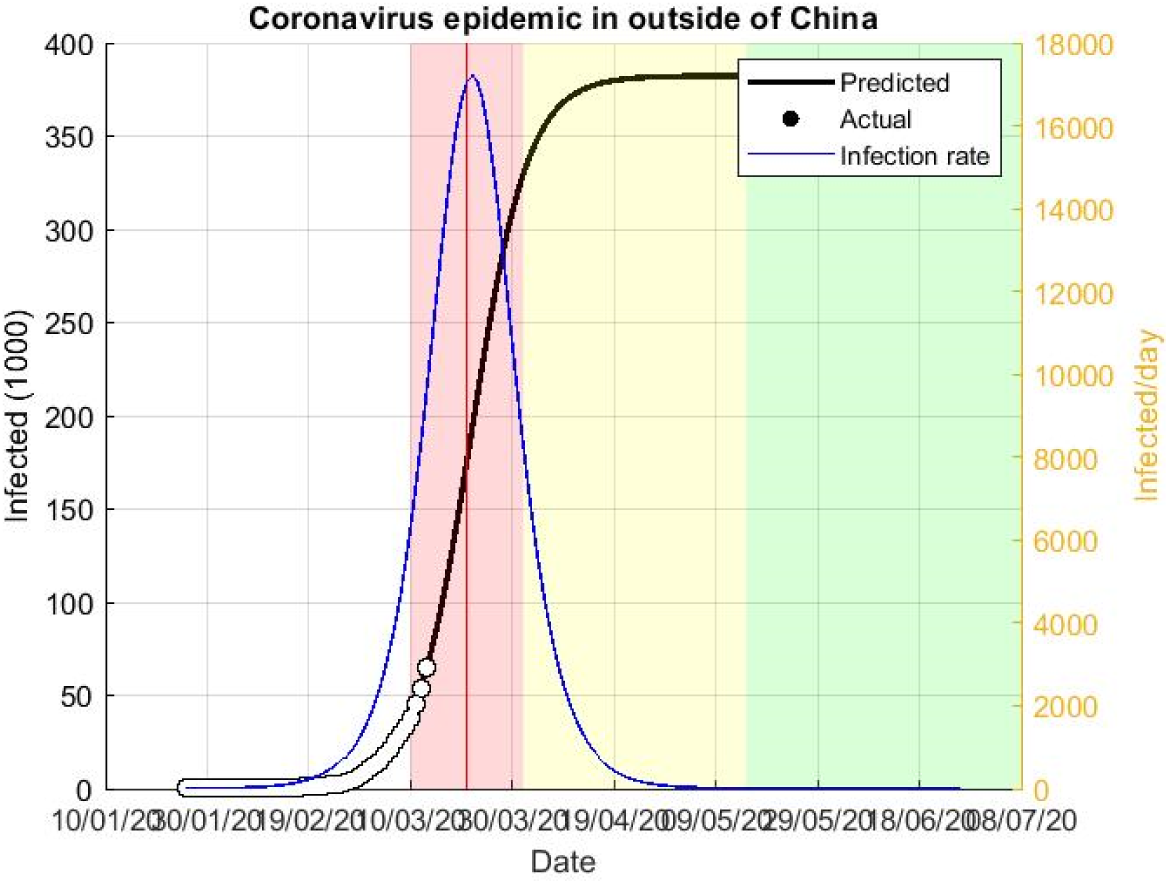
Predicted evaluation of coronavirus epidemic outside of China.

**Figure 7.**
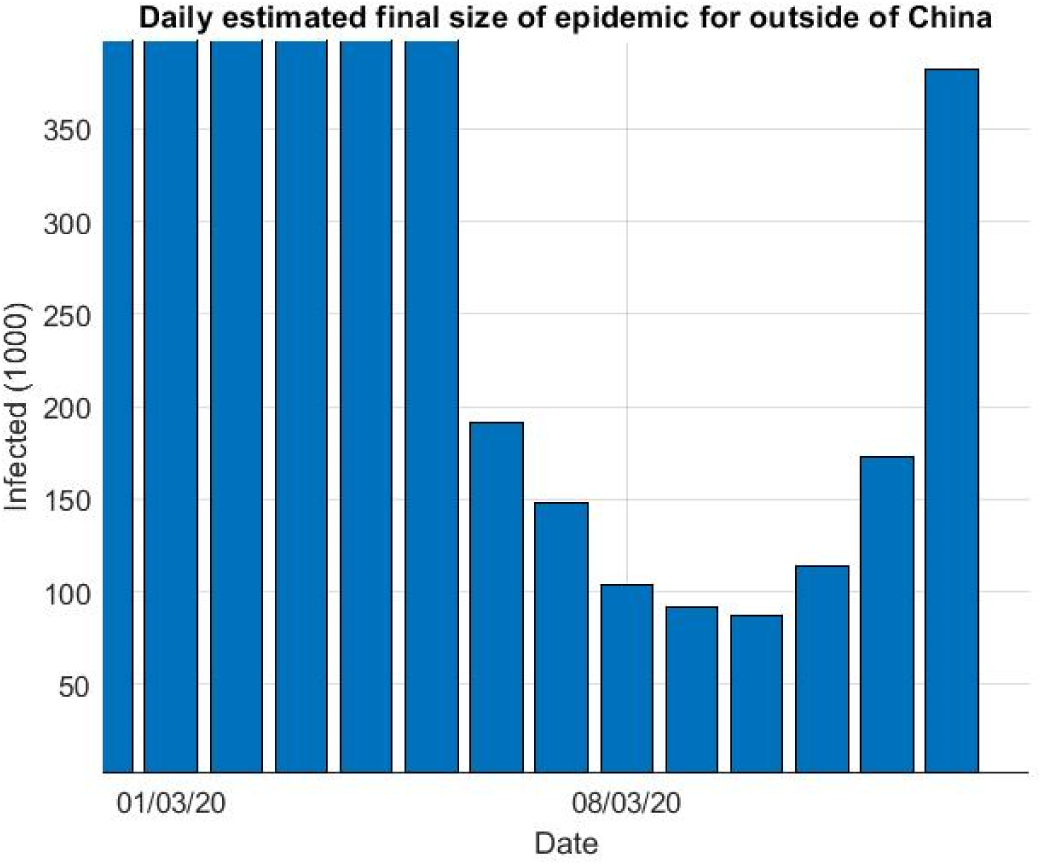
Predicted final size of coronavirus epidemic outside of China.

## 4 Conclusion

On the base of available data, one can predict that the final size of coronavirus epidemy in China will be around 81 000 cases. For South Korea, a prediction is about 8000 cases. For the rest of the World, the forecasts are still very unreliable in is now approximately 380 000 cases.

We emphasize that the logistics model is a phenomenological, data-driven model. Thus, its forecasts are as reliable as useful are data and as good as the model can capture the dynamics of the epidemic. As daily epidemic size forecasts begin to converge, it can be said that the outbreak is under control. However, any systematic deviation from the forecast curve may indicate that the epidemic is escaping control. An example is China. By 25. Feb., the data follows a logistic curve and then begins to deviate from it. We now know that this was the beginning of the second stage of the epidemic, which is now spreading around the World. A similar linear trend can now be observed for South Korea (Fig 5); we hope this does not mark the beginning of the second stage of the epidemic in this country.

## Data Availability

Data are freely available on the net.

https://www.worldometers.info/coronavirus/

## References

[1] M. Batista, Estimation of the final size of the COVID-19 epidemic, medRxiv, (2020) 2020.2002.2016.20023606.

[2] X.S. Wang, J.H. Wu, Y. Yang, Richards model revisited: Validation by and application to infection dynamics, J Theor Biol, 313 (2012) 12–19.

[3] B. Pell, Y. Kuang, C. Viboud, G. Chowell, Using phenomenological models for forecasting the 2015 Ebola challenge, Epidemics, 22 (2018) 62–70.

[4] F. Brauer, Mathematical epidemiology: Past, present, and future, Infectious Disease Modelling, 2 (2017) 113–127.

[5] S.L. Chowell G, Viboud C, Kuang Y., West Africa Approaching a Catastrophic Phase or is the 2014 Ebola Epidemic Slowing Down? Different Models Yield Different Answers for Liberia., PLOS Currents Outbreaks., (2014).

[6] C.M. Bender, S.A. Orszag, Advanced mathematical methods for scientists and engineers I asymptotic methods and perturbation theory, Springer, New York, 1999.

[7] P.F. Verhulst, Notice sur la loi que la population poursuit dans son accroissement, Correspondance mathématique et physique, 10 (1838) 113–121.

